# Development and community evaluation of continuous pressure monitoring for pressure ulcer prevention and management: protocol for a mixed-methods co-design programme

**DOI:** 10.64898/2026.11.18.26367585

**Authors:** Peter R. Worsley, Nicci Aylward-Wotton, Rashedul Hoque, Silvia Caggiari, Euan Sadler, Bridie Kent, Liudi Jiang, Davide Filingeri, Stephen Duckworth, Rebecca Hardwick, Joanne Pendray, Heather Hopper, Reg Taylor, Cynthia Calton, Caroline Searight, Joanne Lloyd, Vanessa Mason, Lynn Tatnell

## Abstract

Pressure ulcers cause substantial preventable harm, reduce quality of life and increased health and care costs. Continuous pressure monitoring (CPM) provides longitudinal information on interface pressure, posture and mobility, but evidence remains limited on its implementation, design requirements, safety and acceptability in community care. The overall aim is to develop and evaluate a theory-informed, co-produced CPM system and implementation strategy for pressure ulcer prevention and management in community settings.

This sequential, theory-informed programme will comprise three linked work packages and a supporting technical-development strand. Work package 1 will use realist evaluation involving focus groups, observations and interviews to develop and refine theories explaining how CPM works, for whom and under which community circumstances. Work package 2 will use appreciative inquiry to co-produce design and implementation requirements. Technical development will integrate these requirements with mobility and pressure exposure analytics. Work package 3 is a nine-month, multisite observational mixed-methods evaluation involving 60 adults at risk of, or living with, pressure ulcers and approximately 20 carers, professionals and NHS staff across private homes, care homes and nursing homes. Outcomes will include usability, user experience, technology acceptance, feasibility, implementation, posture and mobility, pressure exposure, skin status, health-related quality of life and safety.

The University of Southampton sponsors the programme, with approvals governed through the relevant UK Health Research Authority and research ethics processes. Findings will be disseminated through peer-reviewed publications, conference presentations, stakeholder networks, accessible summaries and patient and public involvement and engagement activities.

This multi-site community evaluation will link evidence synthesis, programme theory, co-design, technical analytics and human-factors evaluation, to identify the conditions required for CPM to be usable, acceptable and feasible in community practice, creating the basis for a future definitive evaluation.

## Introduction

### Background

Pressure ulcers (PUs) are localised injuries to the skin and underlying tissue caused by sustained pressure or pressure combined with shear (1). They remain common among people whose ability to reposition is reduced by frailty, neurological impairment, illness, pain, sedation or dependence on others. Their persistence despite national and international guidelines exposes the limitations of periodic risk scores, generic repositioning schedules and labour-intensive care (1, 2) . PUs cause pain, sleep disruption, loss of independence, social isolation and reduced wellbeing (3), while prevention and treatment require substantial clinical resources. UK estimates indicate a major and increasing cost of chronic wounds to the National Health Service (4), although expenditure does not ensure timely, individualised or effective prevention.

Prevention centres on risk assessment, skin inspection, pressure redistribution and repositioning, but each has limitations. Risk tools provide intermittent estimates and have uncertain effects on incidence (5, 6); skin inspection may detect visible damage only after underlying processes have begun (7); support surfaces redistribute rather than minimize loading (8, 9); and prescribed repositioning does not confirm effective off-loading (10). In community care, brief clinical contact, variable staffing, fragmented responsibility and reliance on patients or carers further exacerbate these limitations. There is a need for technology to compliment practice to improve personalisation and effectiveness. An example for pressure ulcer prevention is conventional pressure mapping which provides a short snapshot of interface pressures to aid with support surface selection and self-awareness of pressures. In recent years, these devices have been modified to monitor over prolonged periods, revealing sustained loading, posture and movement over time (11-13), therefore they can be referred as continuous pressure monitoring (CPM) systems. However, measurement alone is far from effective prevention: users must identify clinically meaningful signals and translate them into feasible and sustained action.

Pressure mapping supports cushion prescription and repositioning, but interpretation is operator dependent and brief assessments may not represent daily exposure (14, 15). Continuous systems extend observation but may introduce artefact, data loss, placement errors, installation burden and data-governance requirements (16), and may alter attention and decision-making (17). Although prolonged pressure data can identify posture, mobility and sustained exposure (11, 18-20), evidence of reduced interface pressure remains a surrogate for clinical benefit (16). CPM could therefore augment rather than replace clinical judgement, could support staff resource allocation, appropriate equipment and person-centred care.

### Current evidence and knowledge gap

A recent scoping review identified 24 CPM studies across eight countries, with 67% of them being hospital based (16). Some studies reported improved recognition of loading, repositioning, staff confidence or patient engagement, but many used small samples, short monitoring periods, surrogate outcomes or self-report. Heterogeneity and risk of bias limit comparison and causal inference, and pressure ulcer incidence was not consistently reduced. Technical accuracy or favourable usability does not establish clinical utility, cost-effectiveness, equitable access or sustainable implementation. Research must therefore specify how data are expected to change decisions and outcomes, identify where that pathway fails, and assess burdens and unintended effects.

Prior evaluation of CPM implementation has been completed through Quality Improvement projects, including the ‘Pressure Reduction through Continuous Monitoring in the Community Setting’ (PROMISE) project (21). This was a multisite quality-improvement initiative that monitored 107 patients across four community adopter sites (21). Reported benefits included greater awareness, improved decision-making and equipment selection, and better patient and practitioner experience. However, the design did not isolate CPM effects from concurrent clinical input, equipment changes or increased attention. Systems could also be cumbersome, costly, technically fragile and difficult to install, transfer and interpret. These barriers may affect access, workload, data completeness and continued use, potentially requiring redesign of both the device and its service model.

Parallel work addressed how high-volume pressure maps could become clinically interpretable. A secondary PROMISE analysis combined buttock-region pressure magnitude with exposure duration using a sigmoid pressure–time relationship (17). Alongside earlier machine-learning studies (11, 18, 19), it established technical feasibility. However, algorithmic classification depends on sensor data quality, anatomical coverage, modelling assumptions, thresholds and the uncertain relationship between interface pressure and tissue risk (22). Retrospective discrimination does not establish prospective validity, generalisability or patient benefit. Evaluation must therefore determine whether outputs are trustworthy, actionable and proportionate to workload and the risk of false reassurance or unnecessary intervention. The remaining knowledge gap is therefore whether CPM can be integrated into diverse community work systems without transferring disproportionate burden or risk to patients, carers and professionals.

### Rationale for the research programme

Technical performance alone will not determine benefit: CPM must fit physical environments, workflows and digital infrastructure, and users must understand and act on its outputs. The SEAMLESS programme outlined in this protocol manuscript will therefore use an iterative evidence-to-implementation model (Figure 1). Evidence synthesis and realist evaluation will form a co-development cycle linking programme theory, stakeholder requirements and engineering feasibility (Work Packages 1 and 2). The final evaluation (Work Package 3) will evaluate the integrated system and inform refinement before a definitive evaluation. This will avoid treating technology as finished before causal explanations for successful implementation are understood.

**Figure 1.**
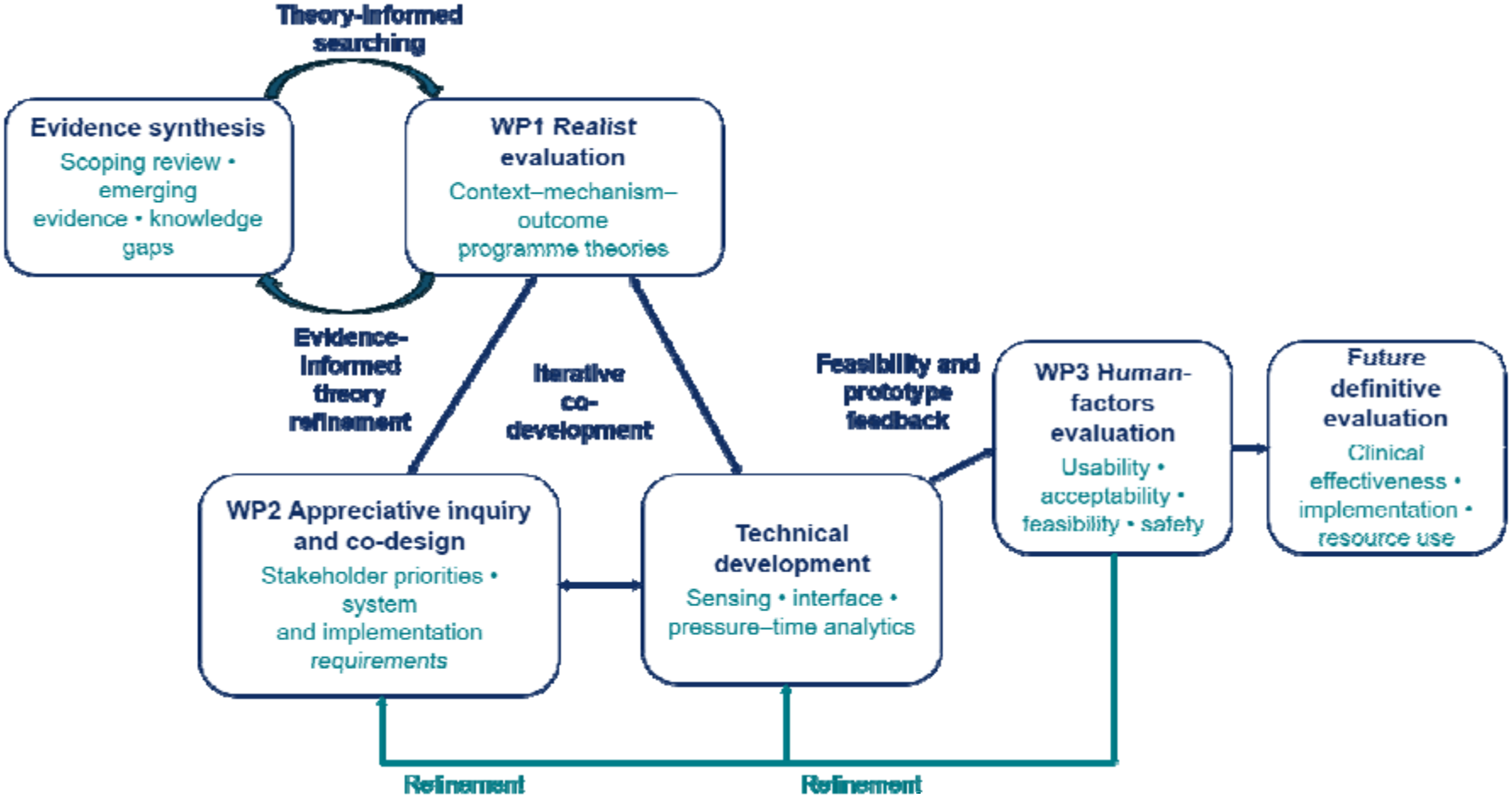
Iterative programme model linking evidence synthesis and realist evaluation with reciprocal co-development across work packages. Human-factors findings will inform further refinement before a future definitive evaluation.

### Aims and research questions

The overall aim is to develop and evaluate a theory-informed, co-produced CPM system and implementation strategy for pressure ulcer prevention and management in community settings.

### Objectives

1. Develop, test and refine programme theories explaining how CPM is used by patients, carers and practitioners across community contexts.
2. Identify the contextual conditions and mechanisms associated with engagement, clinical interpretation, self-management and care decisions.
3. Co-produce design principles, system requirements, training and implementation support for community CPM.
4. Integrate longitudinal pressure data with methods for characterising posture, mobility and pressure exposure.
5. Assess usability, user experience, acceptability, recruitment feasibility, implementation, safety and exploratory patient outcomes in a multisite community evaluation.
6. Generate parameters for a future definitive evaluation of clinical effectiveness and implementation.

### Research questions

1. How, why, for whom and under what circumstances does CPM support pressure ulcer prevention and management in community care?
2. Which features of current CPM systems and implementation practices do stakeholders value, and what should be retained or redesigned?
3. Can a co-produced CPM system and implementation strategy be used effectively, efficiently, safely and acceptably by intended users in private homes, care homes and nursing homes?
4. Can CPM-derived posture, mobility and pressure-exposure measures be collected and interpreted alongside user-reported, clinical and feasibility outcomes?

## Methods and analysis

### Programme design

The study will use a sequential, multiphase mixed-methods design. Work package 1 will generate and refine initial (explanatory) programme theories. Work package 2 will use these theories and stakeholder experience to co-design developments in the technology and tools to facilitate its implementation. A linked technical-development strand will operationalise the sensing, interface and analytical components. Work package 3 will prospectively evaluate the integrated approach in a multisite observational human-factors study. Integration is planned across the design, methods, analysis, interpretation and reporting levels, using connecting, building and merging strategies [23]. Each stage will inform the next while retaining its distinct protocol identifier, sample and analytical plan (Table 1).

**Table 1.** Programme logic.

| **Stage** | **Purpose** | **Participants or evidence** | **Principal methods** | **Output informing the next stage** |
|---|---|---|---|---|
| **Evidence synthesis** | Define the clinical and methodological evidence gap | Published clinical CPM literature | Scoping review | Evidence domains, limitations and research priorities |
| **WP1: realist evaluation** | Explain how CPM works, for whom, in which circumstances and why | Patients, carers and community professionals | Realist evaluation, focus groups, observations and interviews | Refined programme theories and context–mechanism–outcome configurations |
| **WP2: appreciative inquiry and co-design** | Translate programme theory and experience into system and implementation requirements | Patients, carers, public contributors, professionals, academics, service providers, data scientists and industry | Discovery, Dream, Design and Destiny workshop cycle | Agreed design specification, training and implementation principles |
| **Technical development** | Operationalise sensing, user-interface and analytical requirements | Co-design outputs and existing pressure datasets | System engineering and pressure–time analytics | Deployable CPM configuration and interpretable outputs |
| **WP3: human-factors evaluation** | Assess usability, experience, feasibility, implementation and safety | 60 patients and approximately 20 other intended users | Multisite mixed-methods evaluation using a realist approach | Parameters and refinements for a future effectiveness study |

The programme is informed by the realist evaluation (23), appreciative inquiry (24), sociotechnical co-design (25) and UK medical-device human-factors guidance (26). In realist terms, context includes the organisational, social, material and personal conditions in which CPM is introduced; mechanisms comprise the resources offered by the intervention and users’ reasoning in response; and outcomes include changes in understanding, behaviour, care decisions, implementation and clinical processes (23). The appreciative inquiry (AI) provides a strengths-based route from explanation to co-design, while human-factors evaluation assesses whether the resulting technology can be used safely, effectively and satisfactorily by intended users in intended environments (26).

## Work package 1: Realist evaluation

### Design and initial programme theories

Work package 1 asks what works, for whom, under which circumstances, how and to what extent. Initial ‘hunches’ derived from PROMISE and the literature (16, 21) will be expressed as if–then propositions and context–mechanism–outcome configurations (Figure 2). Candidate theories include whether visual feedback increases understanding and empowerment; whether prolonged monitoring gives professionals a more accurate account of lifestyle, equipment and postural needs; and whether training enables practitioners to translate CPM data into shared decisions and preventive action.

**Figure 2.**
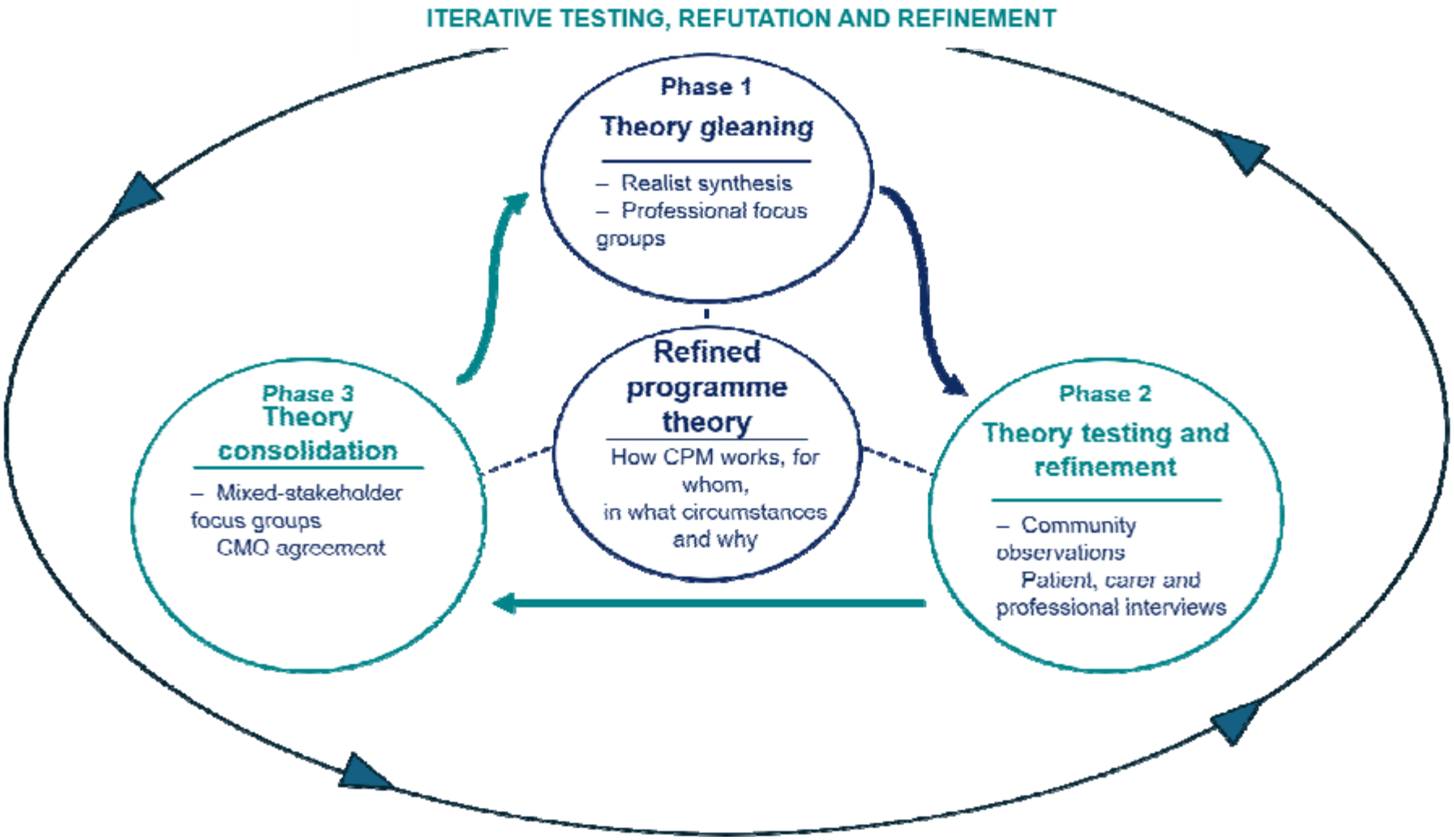
Iterative realist evaluation cycle comprising theory gleaning, theory testing and refinement, and theory consolidation. Evidence from realist synthesis, focus groups, community observations and interviews will be used to develop and agree context–mechanism–outcome configurations and a refined programme theory for CPM.

### Phase 1: Theory gleaning

#### Sampling and recruitment

Participants will be purposively sampled from community teams involved in PROMISE and from patients receiving CPM-supported assessment. Clinical gatekeepers will identify potentially eligible participants and provide approved study information. Interested individuals will contact the research team directly or give permission to be contacted. Sampling will seek variation in dwelling type, pressure ulcer status, mobility, care arrangement and prior CPM experience. The anticipated sample comprises approximately 15 patients, 10 healthcare workers and 10 paid or unpaid carers. Recruitment and analysis will proceed iteratively so that emerging programme theories can inform subsequent sampling and questioning.

Up to four focus groups will be conducted with professionals from community teams experienced in CPM. Each participant will attend one group of up to 60 minutes, in person or online. Discussion will examine current use, challenge or refine the initial propositions, and identify additional contextual influences, resources, reasoning and outcomes. A targeted realist synthesis will run alongside stakeholder consultation, drawing on literature concerning pressure ulcer prevention, biofeedback, repositioning, sensing arrays, interface pressure, artificial intelligence and machine learning.

### Phase 2: Theory testing and refinement

Approximately 15 episodes of CPM use will be observed across private homes, residential care homes and nursing homes in Cornwall, Devon and Somerset. Maximum-variation sampling will seek diversity in care context, patient need and provider. Researchers will record field notes and use a structured observation guide to examine setup, interaction, interpretation, communication and care decisions without interfering with care.

Semi-structured interviews will be conducted with participating patients, carers and healthcare workers, normally between 48 hours and two weeks after observation. Interviews will last up to 60 minutes, be audio-recorded with consent to test and refine initial programme theories following the “gleaning” phase and explore understanding, empowerment, experience, workflow, prior use, decision-making and implementation factors (27). Questions will be adapted iteratively as theories are supported, refuted or elaborated.

### Phase 3: Theory consolidation

Up to two mixed-stakeholder focus groups, each comprising approximately 6–10 patients, carers and professionals, will review candidate context–mechanism–outcome configurations. Groups will seek agreement on how CPM produces outcomes across settings and identify residual disagreements or boundary conditions. The main output will be several refined programme theories for CPM-supported pressure ulcer prevention and treatment in community care.

### WP1 analysis

Interview transcripts will be analysed alongside field notes, observation records and the developing evidence synthesis. Coding will first examine initial programme theories and categories of context, resource, reasoning and outcome, then capture unanticipated explanations and counterexamples. Retroductive reasoning will identify mechanisms that could account for outcomes in particular contexts (28, 29). Configurations will be compared across groups and settings, reformulated and tested in subsequent iterative data collection. NVivo will support data organisation. At least two researchers will discuss coding, rival explanations and negative cases; the wider team and stakeholders will challenge interpretations. A reflexive log will document assumptions, decisions and prior involvement in PROMISE.

### Work package 2: Appreciative inquiry and co-design

#### Design

Work package 2 will use the four-stage appreciative inquiry cycle (30). Discovery will identify valued features of current practice; Dream will explore desirable future CPM; Design will translate ideas into feasible requirements; and Destiny will agree priorities and commitments for implementation. The approach will be strengths based while retaining explicit consideration of limitations, safety and feasibility.

#### Participants and procedures

Up to 40 participants will be purposively selected to achieve representation from patients, paid and unpaid carers, public contributors, tissue-viability and community clinicians, academics, health and social care providers, data scientists, digital and information-governance staff, procurement specialists and industry. Participants will attend a two-day in-person workshop facilitated independently of the core research team. The Discovery phase will identify occasions when current CPM has worked well and the features, relationships and resources that enabled success. The Dream phase will invite participants to describe an ideal future system without initially constraining ideas to the current product. During Design, mixed groups will convert valued features and ambitions into feasible requirements for hardware, software, data presentation, training, cleaning, transport, installation, maintenance and service integration. During Destiny, participants will prioritise requirements, identify responsibilities and agree which elements should progress to development and evaluation.

A subsequent half-day online meeting will present a structured summary of the workshop outputs. Participants will be invited to confirm, amend or reject the proposed requirements and to identify implications that may have been overlooked. Written comments will also be accepted from people unable to attend. Agreement will not be defined as simple unanimity: the team will record the degree of convergence, retain materially different views and document trade-offs between desirability, technical feasibility, affordability, information governance and safety.

### WP2 analysis and output

Workshop recordings, table notes, observation records, individual reflections and prioritisation outputs will be collated into a design matrix. Requirements will be grouped under physical system, user interface, analytical feedback, interoperability and data governance, training, installation, maintenance and service integration. The research team, facilitators and public contributors will compare outputs across stakeholder groups, identify convergence and disagreement, and map each requirement to the WP1 programme theories it is intended to enable. An audit trail will record the source, rationale, feasibility judgement and final status of each requirement. The output will be a version-controlled specification and implementation package for technical development and WP3. During the AI Workshop, members of the research team will independently undertake an Equality and Health Inequalities Impact Assessment (EHIA) using the NHS England EHIA template (31). In addition, participation and evaluation of co-design principles will be assessed by a combined team of research and PPIE members, using the defined principles for co-production (32).

### Technical-development strand

Technical development will link the co-produced requirements to a deployable CPM system. The system will comprise a thin pressure sensing array placed on a bed or chair, a monitor and user interface, secure data handling, and analytical outputs intended to support interpretation and action. Training materials and implementation support will be refined alongside the technology.

The analytical pipeline will characterise static postures, mobility events and pressure exposure. A previously published secondary analysis combined pressure magnitude at the buttock region with exposure duration and a sigmoid pressure–time relationship to categorise exposure risk (18). The technical strand will use this prior work from PROMISE and CPM data obtained from the realist evaluation to inform processing and presentation of parameters which could provide feedback to clinicians, patients and carers.

### Work package 3: Multisite human-factors evaluation

#### Design and setting

Work package 3 is a nine-month observational study of CPM usability and user experience when delivered as part of usual care. It will be conducted across community healthcare services in 4 community NHS care providers. Intended-use environments will include private homes, care homes and nursing homes.

#### Participants

The planned sample comprises 80 participants: 60 adult patients and approximately 20 carers, health and social care professionals, and NHS staff involved in digital systems, information governance or procurement. Eligible patients will be aged 18 years or older; live in a community setting; be at risk of, or have, a pressure ulcer; receive preventive or treatment interventions; and be likely to spend more than eight hours in bed within a 24-hour period. They must be able to provide informed consent or participate through an appropriate representative where capacity is lacking. Carers and professionals must have relevant experience of observing, using or supporting CPM. Patients younger than 18 years, those not at risk of pressure ulceration and those unable to participate through consent or an appropriate representative will be excluded. Participants will be recruited through participating clinical teams using convenience sampling, with efforts to include variation in age, sex, ethnicity and community setting. A delegated investigator will confirm eligibility before enrolment, with criteria illustrated in Table 3. Reimbursement will follow the approved protocol.

### Consent and participation where capacity is impaired

Written informed consent will be obtained in accordance with Good Clinical Practice and the approved protocol. Where a patient lacks capacity, an appropriate personal or professional legal representative process will be used. If capacity is regained, consent for continued participation will be sought. Participation is voluntary, questions may be asked at any time, and withdrawal will not affect care.

### CPM assessment

A commercially available, appropriately marked CPM system will be used in accordance with manufacturer instructions and local care procedures. This consists in a thin flexible sensing mat which will be placed beneath a sheet or suitable cover on a bed, or used on a chair, to record interface pressure distributions. Bed monitoring is expected to continue for up to 72 hours and chair monitoring for approximately one day, according to clinical need and feasibility. Visual and analytical information may inform support-surface selection, repositioning, posture and mobility advice as part of usual care.

Before deployment, participating staff will receive standardised training covering eligibility, infection prevention, sensor placement, system checks, troubleshooting, secure data handling, interpretation of the display and escalation of clinical or technical concerns. Training will be available through face-to-face instruction, written manuals and online materials. The study will record who installs and removes the system, training completed, time required, deviations from intended use, technical support requested and reasons for incomplete monitoring. These fidelity data will distinguish problems inherent to the device from problems arising through training, workflow or local infrastructure.

### Baseline and process data

Baseline variables will include age, sex, ethnicity, height, weight, comorbidities, medication, smoking and alcohol status, continence, skin and wound history, pressure ulcer risk, and current preventive interventions. Process measures will include recruitment, retention, missing data, staff time, training, installation, technical problems, protocol deviations, adverse events and the completeness of CPM recordings.

### Outcomes

#### Primary outcome

Usability and user experience of implementing CPM for pressure ulcer prevention or management in community settings. Usability will be characterised through validated questionnaire scores (System Usability Score) (33), observed or reported use problems, installation and completion data, and qualitative accounts from patients, carers and professionals. The study is designed to identify whether intended users can deploy and interact with the system effectively and with acceptable effort, rather than to declare effectiveness from a single numerical threshold.

#### Secondary outcomes

feasibility of identifying, approaching and recruiting eligible participants; recruitment and attrition; completeness of CPM and questionnaire data; acceptability (Technology Assessment Model) (34) and intention to use; implementation barriers and facilitators; staff time and support requirements; posture, mobility, pressure magnitude and exposure duration; baseline and follow-up skin status where available; health-related quality of life (EQ-5D) (35); technical faults; protocol deviations; and adverse events. These findings will inform progression criteria for a future study, including recruitment, retention, data completeness, deployability and the absence of unresolved safety concerns. Numerical thresholds will be agreed with the steering group before outcome data are reviewed.

### Qualitative data collection

Face-to-face or remote semi-structured interviews of up to 60 minutes will explore usability, user experience, perceived value, training, confidence, workflow, clinical integration, self-management, implementation barriers and facilitators, and willingness to continue using the system. Interviews will be audio-recorded with consent and transcribed verbatim.

### Analysis and integration

#### Quantitative analysis

Categorical variables will be reported as counts and percentages with explicit denominators. Continuous variables will be examined graphically and summarised as means with standard deviations or medians with interquartile ranges, as appropriate; 95% confidence intervals will be presented where informative. Recruitment, consent, retention and completion will be summarised as counts and proportions with 95% confidence intervals, and cumulative recruitment will be plotted by time and site. Questionnaire scores will be calculated according to published instructions and summarised overall and by respondent group. Exploratory comparisons may examine setting, previous CPM experience and participant characteristics where sample sizes permit. Any pre–post or subgroup analyses will be explicitly exploratory and interpreted using estimates and uncertainty rather than significance alone.

#### Qualitative analysis

Interview transcripts will be analysed using realist thematic analysis (36). Researchers will familiarise themselves with the data, generate initial codes, develop and refine themes, examine patterns across user groups and settings, and produce an interpretive account of system use. Analysis will focus on usability problems, perceived usefulness, confidence, training, workflow, burden, communication, responses to CPM feedback and willingness to continue use. Contradictory and less favourable accounts will be retained. A coding framework and analytic memos will provide an audit trail, while discussion within the qualitative team and with PPIE contributors will broaden interpretation rather than seek mechanical inter-rater agreement.

#### Mixed-methods integration

Integration will occur across design, methods, analysis, interpretation and reporting (37). Findings from each component will shape subsequent questions, sampling and procedures. In the human factors trial, quantitative, qualitative, process and technical datasets will be analysed separately and then merged in participant-, setting- and theory-level joint displays (38). Displays will align questionnaire and CPM measures with installation data, incidents and interview themes and generate meta-inferences beyond either dataset alone. Convergence coding will record agreement, complementarity, expansion and dissonance (39); discordant findings will be examined as possible explanations of contextual variation. Integrated findings will refine programme theory and inform a future effectiveness and implementation evaluation.

#### Sample-size rationale

WP1 and WP2 will use purposive, information-rich sampling appropriate to realist evaluation and co-production, with iterative recruitment guided by theory refinement and stakeholder diversity. WP3 will aim to recruit 60 patient users and approximately 20 other users. This pragmatic sample is intended to expose a broad range of usability issues and estimate feasibility parameters; it is not powered to detect differences in pressure ulcer incidence.

#### Data management and confidentiality

Participants will be assigned coded identifiers. Consent and contact information will be stored separately from research data. Paper records will be secured at participating organisations, and electronic data will be stored on password-protected institutional systems with role-based access. Audio files will be transferred securely for approved transcription, anonymised during transcription, and deleted or retained according to the approved data-management plan. CPM files will be transferred to secure University of Southampton servers for analysis. Data will be retained for the period specified by the sponsor; the work package 3 protocol specifies a minimum of 15 years. Access to the final dataset and any data sharing will comply with participant consent, UK data-protection legislation, sponsor policy and safeguards for potentially identifiable qualitative data.

#### Patient and public involvement and engagement

Patient and public contributors have shaped the research questions, study design, participant-facing materials and co-production strategy. A standing PPIE group has reviewed information and consent materials and reports to programme governance. Contributors will participate in work package 2, interpretation of anonymised findings where approved, dissemination and development of accessible summaries. Reporting will describe the aims, methods, outcomes and influence of involvement, including positive and negative effects and critical reflection on the process.

#### Ethics, safety and governance

The University of Southampton is the study sponsor. Work packages 1–2 are associated with IRAS 323530. Work package 3 is associated with IRAS 350350 and sponsor reference ERGO2 104003. Before submission, this section will be completed with the approving Research Ethics Committee, reference number, approval date and status for each component, verified against sponsor documentation.

Potential burdens include distress when discussing pressure ulcer experiences, intrusion during observations, fatigue, confidentiality risks and risks associated with research in private homes. Participants may pause or stop an observation, interview, focus group or monitoring episode without giving a reason and without affecting care. If a participant withdraws, the research team will document whether data already collected may continue to be used, in accordance with the relevant consent materials and approved protocol. Researchers will follow University and participating-service lone-working, safeguarding and escalation procedures.

For WP3, any untoward medical occurrence temporally associated with CPM or study procedures will be recorded with onset, resolution, severity, causality, action and outcome. Serious or unexpected events and device incidents will be reviewed by the chief investigator and reported to the sponsor and relevant authorities within applicable timelines. These risks will be mitigated through manufacturer instructions, cleaning procedures, staff training, system checks and retention of clinical responsibility by the treating team. Substantial and non-substantial protocol amendments will be classified by the sponsor and implemented only after required approvals. Deviations will be documented, recurrent deviations investigated, and annual and end-of-study reports submitted as required.

The research team includes members involved in earlier CPM implementation. Reflexivity, multidisciplinary interpretation, explicit testing of rival explanations and stakeholder validation will be used to mitigate confirmation bias. Industry participation in co-design will be declared and managed transparently.

#### Dissemination

Results will be reported to the sponsor and relevant ethics and regulatory bodies and submitted to peer-reviewed journals and scientific conferences. Participants and community partners will receive accessible summaries, and findings will be shared through project, professional and PPIE networks. Publications will distinguish planned from achieved recruitment, report protocol deviations and follow applicable reporting guidance. Authorship will meet journal criteria, with contributions described using the CRediT taxonomy.

## Discussion

This programme will address the gap between recording interface pressure and using that information reliably in community care. CPM can identify loading, posture and mobility and prompt changes to repositioning or equipment (12, 21), but heterogeneous evidence does not show consistent clinical benefit (16). The programme will therefore evaluate the context, mechanisms and implementation processes required for effective use.

The programme responds to these implementation risks by using co-design and human-factors engineering as complementary forms of inquiry. Co-design will identify what patients, carers and professionals value and how responsibilities and trade-offs should be managed; human-factors evaluation will test whether safety-critical tasks can be completed under realistic conditions (25, 40). Their combined value lies in examining the device within its care pathway, professional work, training, technical support, governance and organisational infrastructure rather than assessing usability in isolation (26).

The programme’s principal contribution is a traceable pathway from explanation to design and evaluation. Realist evaluation will identify how setting, professional preparation, patient capability, care relationships and service organisation shape responses to CPM. Those explanations will be translated into explicit system and implementation requirements, then tested against process, technical, quantitative and qualitative evidence before progression to a larger trial. The technical strand is embedded within the programme. Published algorithms combine pressure magnitude and duration and detect posture and mobility (18-20), but their value depends on whether outputs are robust, interpretable and actionable. Mixed-methods integration will examine how data are understood, communicated and incorporated into care decisions.

WP3 is pragmatic and not powered to detect differences in pressure ulcer incidence. Convenience sampling may favour participants receptive to CPM, and researchers’ prior PROMISE involvement may influence interpretation. Maximum-variation sampling, negative-case analysis, reflexivity, multidisciplinary interpretation and retention of discordant accounts will mitigate these risks. The English-language requirement may also limit inclusion and should inform adaptations in a future study. Outputs will comprise a refined programme theory, traceable design and implementation requirements, and estimates of usability, feasibility and safety. These will determine whether, and under which conditions, to progress to a powered evaluation of clinical effectiveness, implementation and resource use.

## Strengths and limitations

- The sequential design will link evidence, explanation, co-production, technical analytics and evaluation rather than treating technology as a standalone intervention.
- Multisite community delivery will capture private-home, care-home and nursing-home contexts and multiple user groups.
- Realist and qualitative methods can explain variation that would be obscured by aggregate usability scores alone.
- The observational design and pragmatic sample are appropriate for human-factors and feasibility objectives but cannot establish effectiveness in preventing pressure ulcers.
- Convenience sampling, previous exposure to CPM and the research team’s involvement in earlier development may influence transferability and responses.
- Participants requiring interpretation are excluded from WP3 under the current protocol, which may limit inclusivity and should be reconsidered in future research.

## Data Availability

All data produced in the present study are available upon reasonable request to the authors

## Submission statements

### Data availability

No datasets were generated or analysed for this protocol article. Data generated by the prospective work packages will be shared with the associated results publications where ethically and legally permissible, subject to participant consent, sponsor policy and safeguards for potentially identifiable qualitative data. Each results publication will provide the relevant repository, persistent identifier and access conditions.

### Author contributions

Contributions will be reported using the CRediT taxonomy once the final author list and order have been agreed.

### Protocol registration

This programme is not a randomised clinical trial. Relevant IRAS, sponsor and research ethics identifiers are reported above. Any prospective public registration record will be added before submission.

### Reporting

The realist component will be reported in accordance with RAMESES II. The submission package will include the PLOS Human Participants Checklist, the approved protocols for work packages 1–2 and work package 3, and the RAMESES II reporting checklist. Subsequent results manuscripts will include the reporting checklists appropriate to their observational, qualitative and mixed-methods designs.

### Figures and Tables

Figure 1. Iterative programme model showing reciprocal evidence synthesis and realist evaluation; cyclical integration of work package 1, work package 2 co-design and technical development; human-factors evaluation and refinement; and progression to a future definitive evaluation.

Figure 2. WP1 realist evaluation cycle showing theory gleaning, theory testing and refinement, and theory consolidation.

Figure 3. WP3 participant pathway and timing of baseline assessment, CPM deployment, questionnaire completion and qualitative interviews.

**Figure 3.**
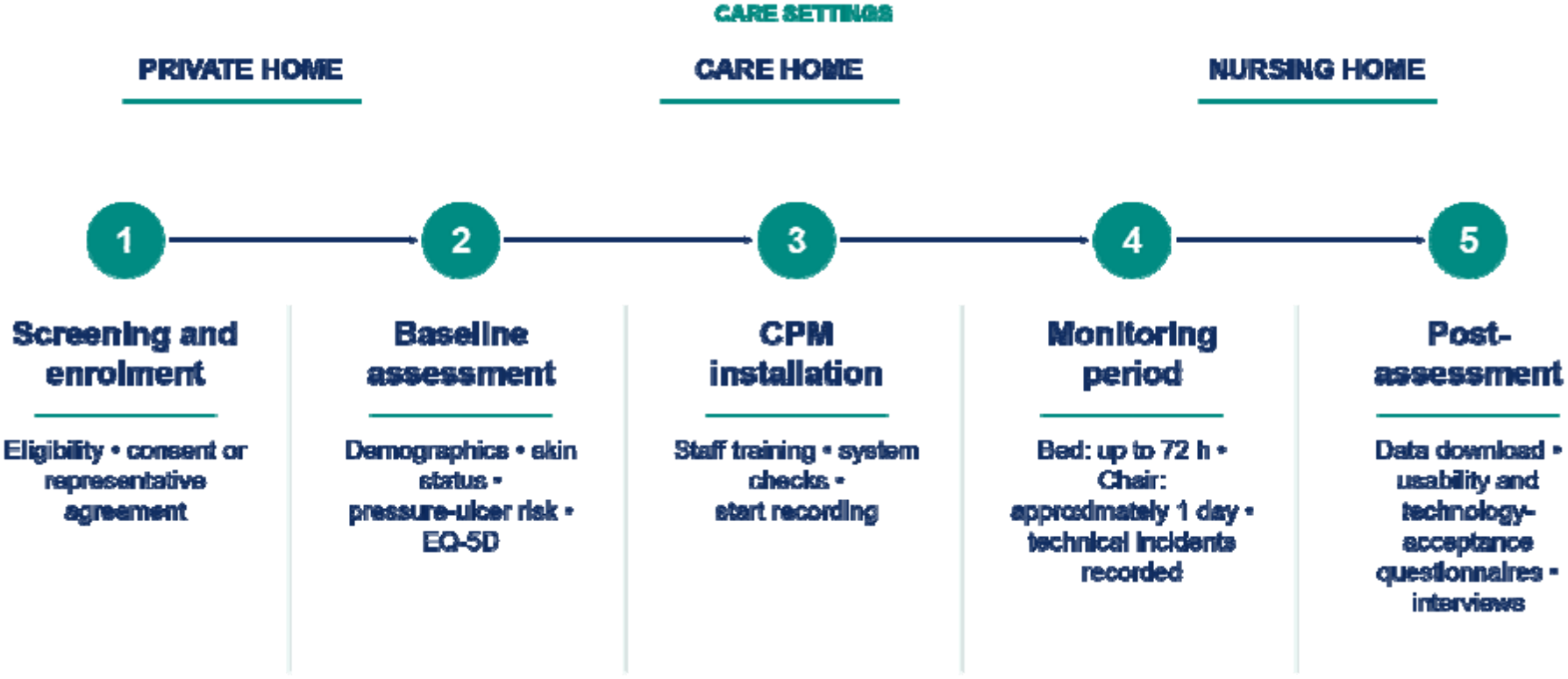
Work package 3 participant pathway across private homes, care homes and nursing homes. Following screening and consent or representative agreement, participants will undergo baseline assessment and CPM installation, monitoring for up to 72 hours in bed or approximately one day in a chair, and post-assessment data download, usability and technology-acceptance questionnaires, and interviews.

Table 1. Sequential programme logic, participants, methods and outputs.

Table 2. WP3 assessment schedule.

**Table 2.** WP3 assessment schedule.

| **Assessment** | **Screening and enrolment** | **Baseline and installation** | **During monitoring** | **After CPM assessment** |
|---|---|---|---|---|
| **Eligibility and consent or representative agreement** | Yes |  |  |  |
| **Demographic and clinical characteristics** |  | Yes |  |  |
| **Skin status, pressure ulcer risk and current prevention** |  | Yes | Clinical care as required | Recorded where available |
| **Installation time, staff input and fidelity** |  | Yes | Technical support and deviations | Removal time and completeness |
| **Continuous pressure, posture and mobility data** |  | Recording starts | Continuous recording | Download and quality check |
| **Usability, acceptance and user experience questionnaires** |  |  |  | Yes, by relevant user group |
| **EQ-5D** |  | Yes |  | Follow-up where specified |
| **Semi-structured interview** |  |  |  | Purposive subsample or relevant participants |
| **Adverse events and technical incidents** |  | From installation | Continuous | Through final contact |

Table 3. Eligibility criteria across prospective programme components.

**Table 3.** Eligibility criteria across prospective programme components.

| **Participant group** | **Inclusion criteria** | **Exclusion criteria or limitations** |
|---|---|---|
| **WP1 patients** | Adult living in a community dwelling; current or previous pressure ulcer, or assessed risk; CPM indicated by the participating clinical team; receiving care from a participating community service. | End-of-life care; unable to participate in observation through an approved consent or consultee process. |
| **WP1 carers** | Adult paid or unpaid carer actively supporting a person at risk of, or living with, a pressure ulcer; has interacted with the patient and professional during CPM. | No experience of the CPM episode under study. |
| **WP1 professionals** | Employed by a participating service; involved in pressure ulcer prevention or treatment; prior use of CPM; consent for the relevant observation, interview or focus group. | No prior CPM use or no active role in pressure ulcer prevention and management. |
| **WP2 stakeholders** | Adult with relevant lived, caring, clinical, academic, technical, service, digital, procurement or industry experience; agrees to workshop participation. | No relevant experience or does not consent to participation. |
| **WP3 patients** | Adult aged 18 years or older; living in a private home, care home or nursing home; at risk of or currently living with a pressure ulcer; receiving preventive or treatment interventions; likely to spend more than eight hours in bed in 24 hours; able to consent or participate through an appropriate representative. | Younger than 18 years; not at pressure ulcer risk; unable to participate through the approved consent or representative process. The current protocol requires sufficient English for the study procedures. |
| **WP3 carers and professionals** | Actively supports pressure ulcer prevention or treatment and has observed, used or supported CPM in care delivery, digital systems, information governance or procurement. | No relevant interaction with CPM. |

